# It’s not TB but what could it be? Abnormalities on chest X-rays taken during the Kenya National Tuberculosis Prevalence Survey

**DOI:** 10.1101/2020.08.19.20177907

**Authors:** Brenda Nyambura Mungai, Elizabeth Joekes, Enos Masini, Angela Obasi, Veronica Manduku, Beatrice Mugi, Jane Onǵanǵo, Dickson Kirathe, Richard Kiplimo, Joseph Sitienei, Rose Oronje, Ben Morton, Stephen Bertel Squire, Peter MacPherson

## Abstract

**Background:** The prevalence of diseases other than tuberculosis (TB) detected during chest X-ray (CXR) screening is unknown in sub-Saharan Africa. This represents a missed opportunity for identification and treatment of potentially significant disease. Our aim was to quantify and characterise non-TB abnormalities identified by TB-focused CXR screening during the 2016 Kenya National TB prevalence survey.

**Methods:** We reviewed a random sample of 1140 adult (≥15 years) CXRs classified as “abnormal, suggestive of TB” or “abnormal other” during field interpretation from the TB Prevalence Survey. Each image was read (blinded to field classification and study radiologist read) by two expert radiologists, with images classified into one of four major anatomical categories and primary radiological diagnosis. A third reader resolved discrepancies. Prevalence and 95% confidence intervals of abnormalities diagnosis were estimated.

**Findings:** Cardiomegaly was the most common non-TB abnormality at 259/1123 (23.1%, 95% CI 20.6%-25.6%), while cardiomegaly with features of cardiac failure occurred in 17/1123 (1.5 %, 95% CI 0.9%-2.4%). We also identified chronic pulmonary pathology including suspected chronic obstructive pulmonary disease in 3.2% (95% CI 2.3%-4.4%) and non-specific patterns in 4.6% (95% CI 3.5%-6.0%). Prevalence of active-TB and severe post-TB lung changes was 3.6% (95% CI 2.6%-4.8%) and 1.4% (95% CI 0.8%-2.3%) respectively.

**Interpretation:** Based on radiological diagnosis, we identified a wide variety of non-TB diagnoses during population-based TB screening. TB prevalence surveys and active case finding activities using mass CXR offer an opportunity to integrate disease screening efforts.

**Funding:** National Institute for Health Research (IMPALA-grant reference 16/136/35).

Evidence before this study
Tuberculosis (TB) remains the leading adult infectious killer in the world. The World Health Organization (WHO) recommends the use of chest X-ray (CXR) as a mass screening tool in TB prevalence surveys and active case finding activities to identify patients eligible for bacteriological investigation. Mathematical modelling suggests that an algorithm incorporating a screening CXR to direct subsequent Xpert MTB/RIF testing is the optimal pathway with lowest number needed to test at acceptable programme costs in active case finding mass screening activities. Increased digital X-ray availability, coupled with the development of Computer Aided Detection (CAD) software for identification of TB, could enable widespread use of CXR in screening for TB in areas with limited access to radiologists. In addition, CXR has an advantage of detecting conditions besides TB. However, the prevalence of diseases other than TB identified by CXR during TB mass screening or TB prevalence survey activities is unknown.
We systematically searched MEDLINE, CINHAL, Global Health and Google scholar databases from 1940–2019 to identify studies that described the prevalence of non-TB CXR findings during TB prevalence surveys or mass screening activities. The WHO Stop TB Department website and reference lists from relevant reviews and studies were used to supplement the search. The search strategy included MESH terms: “Chest Xray” or “Chest-Xray” or “Chest radiograph” or “Mass screening” or “Mass radiography” AND “Tuberculosis screening” or “Tuberculosis triaging” or “TB screening” or “TB triaging” AND “Non-tuberculous” or “Non-TB pathology” or “other pathology”. Our search yielded a number of studies using CXR screening in prevalence surveys as well as active case finding activities. However, studies describing non-TB pathology during mass radiography were few and mostly in the 20^th^ century. A report in Europe between 1946 and 1948 documented 35% non-TB pathology on mass miniature X-rays. Our search did not identify any evidence pertinent to the sub-Saharan African context.

Added value of this study
In this cross-sectional study, we analysed individual-level participant CXR data from the 2016 Kenya National TB Prevalence Survey. Our aim was to quantify and characterise non-TB abnormalities identified by TB-focused CXR screening during the survey. We hypothesised that non-TB abnormalities requiring further clinical review are highly prevalent and need to be considered when implementing CXR screening for TB. Our study identified multiple non-TB diagnoses. The most prevalent was cardiomegaly at 23.1% (95% CI 20.6%-25.6%). We also identified chronic pulmonary pathology including suspected chronic obstructive pulmonary disease (COPD) and non-specific interstitial patterns. Mediastinal masses, excluding goitres, occurred in 0.8% (95% CI 0.4%-1.5%). TB related abnormalities, which may cause chronic respiratory symptoms, such as severe bronchiectasis and/or destroyed lung were present in 1.4%(95% CI 0.8%-2.3%). Median CAD4TB scores were low for the non-TB abnormalities.

Implications of all the available evidence
Our study demonstrated a high prevalence of CXR-identified non-TB abnormalities, including cardiomegaly, chronic pulmonary diseases, post-TB lung disease and non-specific lung diseases. Implementation of CXR TB screening in this context requires detailed health system planning to incorporate provision of care to people with non-TB abnormalities. This could include incorporation of additional tests such as blood pressure monitoring and spirometry as part of community TB screening interventions.

## Introduction

Tuberculosis (TB) remains the leading adult infectious killer in the world.^1^ Despite an estimated nine percent relative increase in case detection in 2018, there are still three million (30%) people with TB who are undiagnosed or not reported to national TB programmes.^1^ In an effort to identify missing people with TB, countries have adopted more sensitive diagnostic tools, including Xpert MTB/RIF, scaled up intensified active case finding (ACF), and adapted their screening and diagnostic algorithms to include chest X-ray (CXR) as a sensitive and efficient high-throughput initial screening test.^2^

Historically, miniature radiography for mass TB screening activities was widely utilized in high-income countries throughout the 20^th^ century.^3–6^ In lower-and middle-income countries (LMIC), however, CXR has been used primarily as a complementary tool to support clinical diagnosis of patients who are sputum smear negative.^7^ Following the findings from national TB prevalence surveys that have employed CXR for screening, there is renewed interest in the utility of CXR for TB screening, and to triage people seeking care with symptoms for further TB investigations.^8–10^ In TB prevalence surveys conducted in LMICs, CXR has shown high sensitivity for pulmonary TB (PTB) (94%, 95% CI 88–98) but poor specificity (73%, 95% CI 68–77), necessitating confirmation with a microbiological test.^11–13^ Mathematical modelling of various TB screening algorithms as well as diagnostic algorithms shows that CXR followed by Xpert MTB/RIF, though resource intensive, has the lowest number needed to screen to identify a case.^14^ Increased digital CXR availability, coupled with the development of computer aided detection (CAD) software for identification of TB, has enabled widespread use of CXR in screening for TB in areas with limited access to radiologists or expert clinicians.^15, 16^

Use of CXR for mass TB screening will identify other conditions, especially those related to the rising burden of non-communicable diseases (NCDs) in LMICs, including cardiovascular disease, chronic respiratory disease and cancer.^17^ A short narrative report from Europe in the 1940s highlighted a significant number 1225/3423 (35%) of non-tuberculous findings in mass radiography screening.^5^ However, there is no contemporaneous evidence about the prevalence of non-TB abnormalities identified during TB prevalence surveys and mass radiographic TB screening interventions. TB prevalence surveys focus on accurate estimation of TB prevalence encouraging intentional over-reading of CXRs for identification of participants eligible for bacteriological testing and not on identifying other abnormalities.^9^ Systematic screening/ACF programs also focus on early detection of active TB.^4^ Computer-Aided Detection for Tuberculosis (CAD4TB) demonstrates high sensitivity for CXR TB diagnosis but is not calibrated for detection of non-TB abnormalities.^18^ Countries are currently adopting CXR screening in combination with CAD software systems, both for mass community TB screening activities, as well as in healthcare settings.^16^ However, the individual and health system implications of the presence of these non-TB CXR abnormalities are unknown.

We therefore set out to characterise and quantify the nature of non-TB abnormalities on abnormal CXRs taken during the 2016 Kenya National TB prevalence survey.^13^ The secondary aim was to calculate CAD4TB v6.0 software analysis scores of the images and compare to expert radiologist diagnosis.^19^ We hypothesized that the use of CXR during TB screening would identify a substantial number of people with non-TB abnormalities who may require further clinical attention.

## Materials and Methods

### Study Design

A cross-sectional study using individual-level participant CXR data from adult community members who took part in the 2016 Kenya National TB Prevalence Survey.^13^ We conducted the study in two parts: an initial pilot study (n = 484) to refine tools and estimate the full sample size required for precision, and the main study (n = 1140). The main aim was to estimate prevalence and uncertainty for CXR-identified non-TB disease pathology within this population.

### Study Population

The Prevalence Survey was a population-based cross-sectional study conducted in 2015-2016. The aim was to determine the prevalence of bacteriologically confirmed PTB among adults (≥15 years) and to assess their health seeking behaviour. The survey used the WHO recommended screening strategy comprising symptom questionnaire and CXR.^9^ There were 63,050 enrolled participants, 62,484 (99%) underwent CXR screening. The survey identified 305 TB cases; weighted national prevalence of 558 [95% CI 455-662] per 100,000 adult population.^12, 13^

### Ethical approval

This study was conducted as part of the Kenya Prevalence survey ethics approval reference number SSC 2094 by Kenya Medical Research Institute. The CXR study used anonymized prevalence survey database and individual consent to participate in this secondary analysis was not required.

### Methods

#### Classification of CXRs during the Kenya TB prevalence survey

Digitally-acquired posterior-anterior CXRs were uploaded to a digital archive. Independent, blinded reading of each film was conducted by two clinical officers in the field who had undergone CXR training. Each image was classified as either: a) normal; b) “abnormal, suggestive of TB”; or c) “abnormal other”. Any participant with a CXR classified as “abnormal, suggestive of TB” by either one of the clinical officers, or with a cough of more than two weeks, was eligible for sputum collection. Those confirmed to have TB were referred for treatment and those with other CXR abnormalities were to be linked to a health facility. The prevalence survey has been reported fully elsewhere.^13^

#### Study procedures

We obtained an anonymised line list of all the participants from Prevalence Survey database; our sampling frame included all CXRs classified as “abnormal, suggestive of TB” or “abnormal other” by the survey field readers. Images selected for inclusion in this study were uploaded to a web-based picture archiving and communication system.

Ten specialist radiologists (five Kenyan, five Global North) with median experience of 12.5 years in TB/chest radiology were recruited (Appendix 1). One-to-one training of the radiologists was provided on the online reporting tool and diagnostic case definitions.^20^ Based on previous reporting tools, we developed radiological diagnostic case definitions, comprised of specific diagnoses within four major anatomical areas (lung parenchyma, heart and great vessels, pleura, and mediastinum).^21^ For each major anatomical area, a list of most common diagnoses was developed, taking into consideration Kenyan disease epidemiology (Appendix 2). Radiologists were able to select one primary diagnosis. Differential diagnoses could be added where a single, confident primary diagnosis could not be made.

Each radiologist was randomly assigned CXRs for review. After completion of each reading, the image was released into a pool for second reading. The readers were blinded to each other’s report, but not to clinical information (sex, age, HIV status and symptoms). Finally, 10% of images were re-allocated to the original readers, for assessment of intra-observer variation. Where pairs of radiologists had discrepant primary diagnoses, one of two additional radiologists undertook a consensus read, with knowledge of the first two radiologists’ classification.

### Data management and analysis

Based on the pilot study findings (Appendix 3), 1140 CXR images (390 “abnormal, suggestive of TB” and 750 “abnormal other”) would be required to estimate the prevalence of cardiomegaly (the most common diagnosis in the pilot study) within 3.5% percentage points of the true value with 95% confidence.

For the main study, we included images classified as either “abnormal, suggestive of TB” and “abnormal other”, excluding those sampled in the pilot study. These images were grouped into 99 strata as per the prevalence survey clusters, and sampled without replacement from each stratum. Statistical analysis used R v3.6.2 (The R Foundation for Statistical Computing, Vienna). Inter- and intra-reader agreement was calculated using the Cohens Kappa statistic. Study participant characteristics were calculated as medians or percentages. The prevalence of primary diagnoses was calculated as the number of CXRs depicting the abnormality divided by the total number of images that were readable; 95% confidence intervals were estimated using the binomial exact method. A number of final diagnoses were combined or removed before analysis, based on their prevalence and likelihood of clinical relevance. The 1140 CXRs were analysed using CAD4TB v6.0 (Delft Imaging Systems, Netherlands).^19^ Median scores and interquartile ranges (IQR) were calculated for each primary diagnostic group.

### Results

Out of 63,050 participants in the Prevalence Survey, 62,484 (99%) underwent CXR, with 50,935 (81.5%) reported as normal by survey field staff, 6,425 (10.3%) as “abnormal, suggestive of TB”, and 5,124 (8.2%) as “abnormal other” (Figure 1).

**Figure 1:**
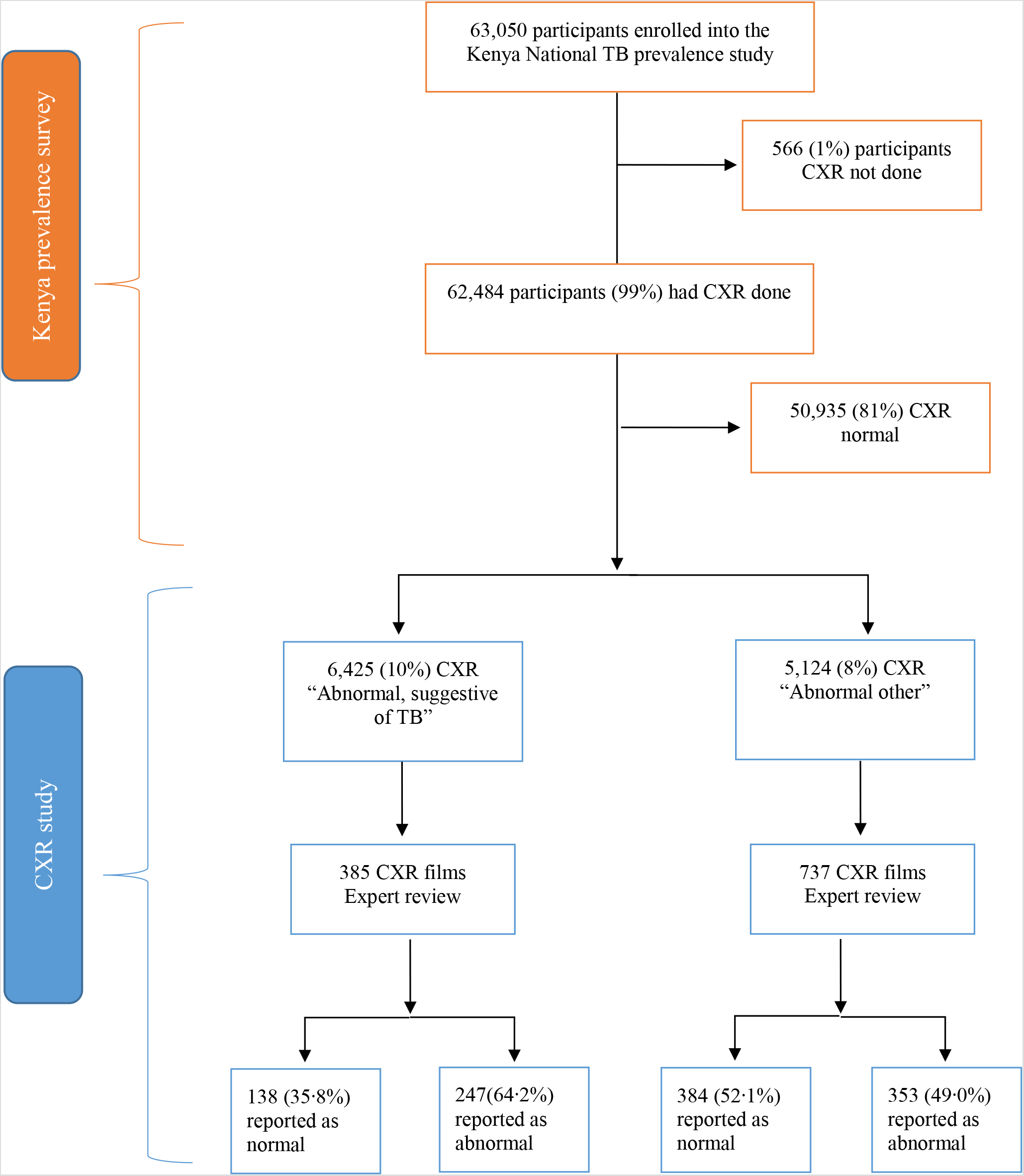
CONSORT Flow Diagram showing participant flow from the Kenya National TB Prevalence Survey and this nested CXR study

#### Participant characteristics

Out of 1140 images included in the main study, 1123 (98.5%) were read by two radiologists and 17 (1.5%) were classified as unreadable. The median patient age was 51.0 years (IQR 36.5–66.0) and 720 (64.2%) were female. Two-hundred and fifty (22.3%) had reported cough, 40 (3.6%) chest pain and 42 (3.7%) self-reported HIV positive status. Six (0.5%) reported current and 34 (3.0%) reported previous TB treatment. GeneXpert and/or culture results were positive for 27/456 (5.9%) of those that had sputum tested (Table 1).

**Table 1:** Characteristics and sputum microbiology results of study participants, stratified by the original survey classification of the chest X-rays as “abnormal, suggestive of TB”, versus “abnormal other”.

|  | Abnormal, suggestive of TB (N=385) | Abnormal, other (N=737) | Total (N=1123) | P-value |
| --- | --- | --- | --- | --- |
| **Sex** |  |  |  | < 0.001 |
| Missing | 0 | 1 | 1 |  |
| Female | 198 (51.4%) | 570 (70.8%) | 720 (64.2%) |  |
| Male | 187 (48.6%) | 239 (29.2%) | 402 (35.8%) |  |
| **Age (years)** |  |  |  | < 0.001 |
| Median (Q1, Q3) | 45.0 (33.0, 64.0) | 54.0 (38.0, 67.0) | 51.0 (36.2, 66.0) |  |
| **CAD4TB score** |  |  |  | < 0.001 |
| Median (Q1, Q3) | 54.0 (47.0, 65.0) | 49.0 (45.0, 53.0) | 50.0 (45.0, 56.8) |  |
| **Study consensus interpretation** |  |  |  | <0.001 |
| Abnormal CXR | 247 (64.2%) | 353 (49.0%) | 600 (53.4%) |  |
| Normal CXR | 138 (35.8%) | 384 (52.1%) | 522 (46.6%) |  |
| Missing |  |  | 1 |  |
| **Cough of any duration** |  |  |  | 0.002 |
| Missing | 0 | 1 | 1 |  |
| No | 278 (72.2%) | 594 (80.6%) | 872 (77.7%) |  |
| Yes | 107 (27.8%) | 143 (19.4%) | 250 (22.3%) |  |
| **Self-reported HIV status** |  |  |  | 0.005 |
| Missing | 183 | 389 | 572 |  |
| HIV-negative | 180 (89.1%) | 328 (94.3%) | 508 (92.4%) |  |
| HIV-positive | 22 (10.9%) | 20 (5.7%) | 42 (7.6%) |  |
| **Taking TB treatment** |  |  |  | 0.069 |
| Missing | 182 | 340 | 522 |  |
| No | 199 (98.0%) | 395 (99.5%) | 594 (99.0%) |  |
| Yes | 4 (2.0%) | 2 (0.5%) | 6 (1.0%) |  |
| **Previously treated for TB** |  |  |  | < 0.001 |
| Missing | 182 | 403 | 522 |  |
| No | 175 (86.2%) | 391 (98.5%) | 566 (94.3%) |  |
| Yes | 28 (13.8%) | 6 (1.5%) | 34 (5.7%) |  |
| **Sputum Xpert/Culture result** |  |  |  | 0.001 |
| Missing | 66 | 600 | 666 |  |
| Negative | 293 (91.8%) | 185 (99.3%) | 429 (94.1%) |  |
| Positive | 26 (8.2%) | 1 (0.7%) | 27 (5.9%) |  |

#### Inter-reader and intra-reader variability

The overall agreement between pairs of readers was moderate with kappa = 0.41 (Appendix 4). There was perfect intra-reader agreement at kappa = 1.

#### Prevalence of CXR abnormalities

Overall, six-hundred (53.4%) images were classified by study radiologists as having any abnormality. Of the images classified as “abnormal, suggestive of TB” by field interpretation in the survey, 203 (64.9%) were classed by expert reviewers as abnormal, whereas among the “abnormal other” category 397 (49.0%) were abnormal by expert radiologist read (Table 1).

Overall prevalence of abnormalities in the major diagnostic categories were: heart and/or great vessels 26.3% (95% CI 23.7%-28.9%), lung parenchyma 26.1% (95% CI 23.5%-28.8%), pleura 7.6% (95% CI 6.1%-9.3%) and the mediastinum 3% (95% CI 2.1%-4.2%) (Figure 2). Among the 600 abnormal images, 21% (127/600) had multiple abnormalities, cardiomegaly accounted for 259/600, 43.2% (39.2%-47.2%) followed by mild/moderate post-TB lung changes at 85/600, 14.2% (11.5%-17.2%) (Figure 3).

**Figure 2:**
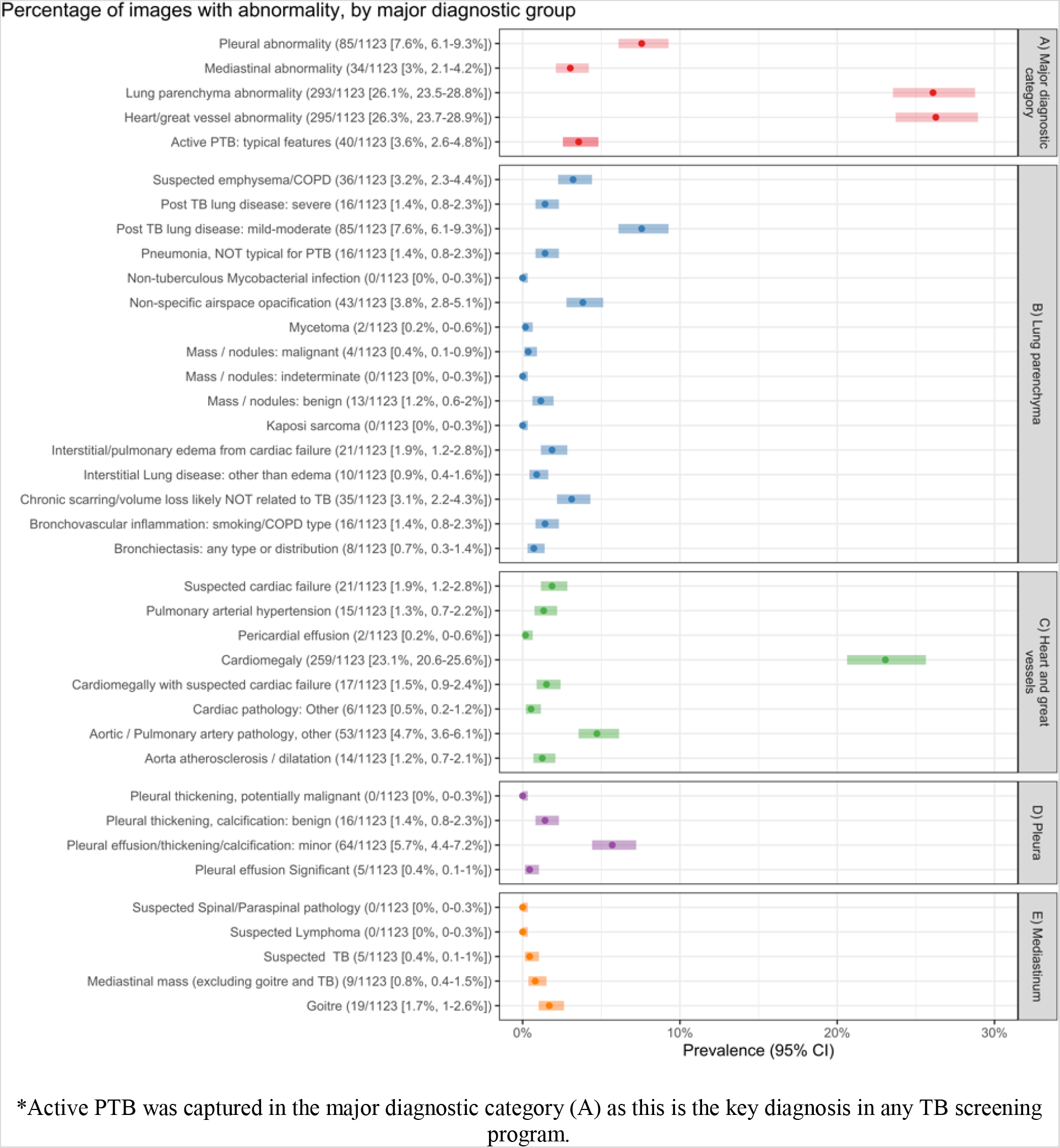
Prevalence of abnormalities by major diagnostic categories

**Figure 3:**
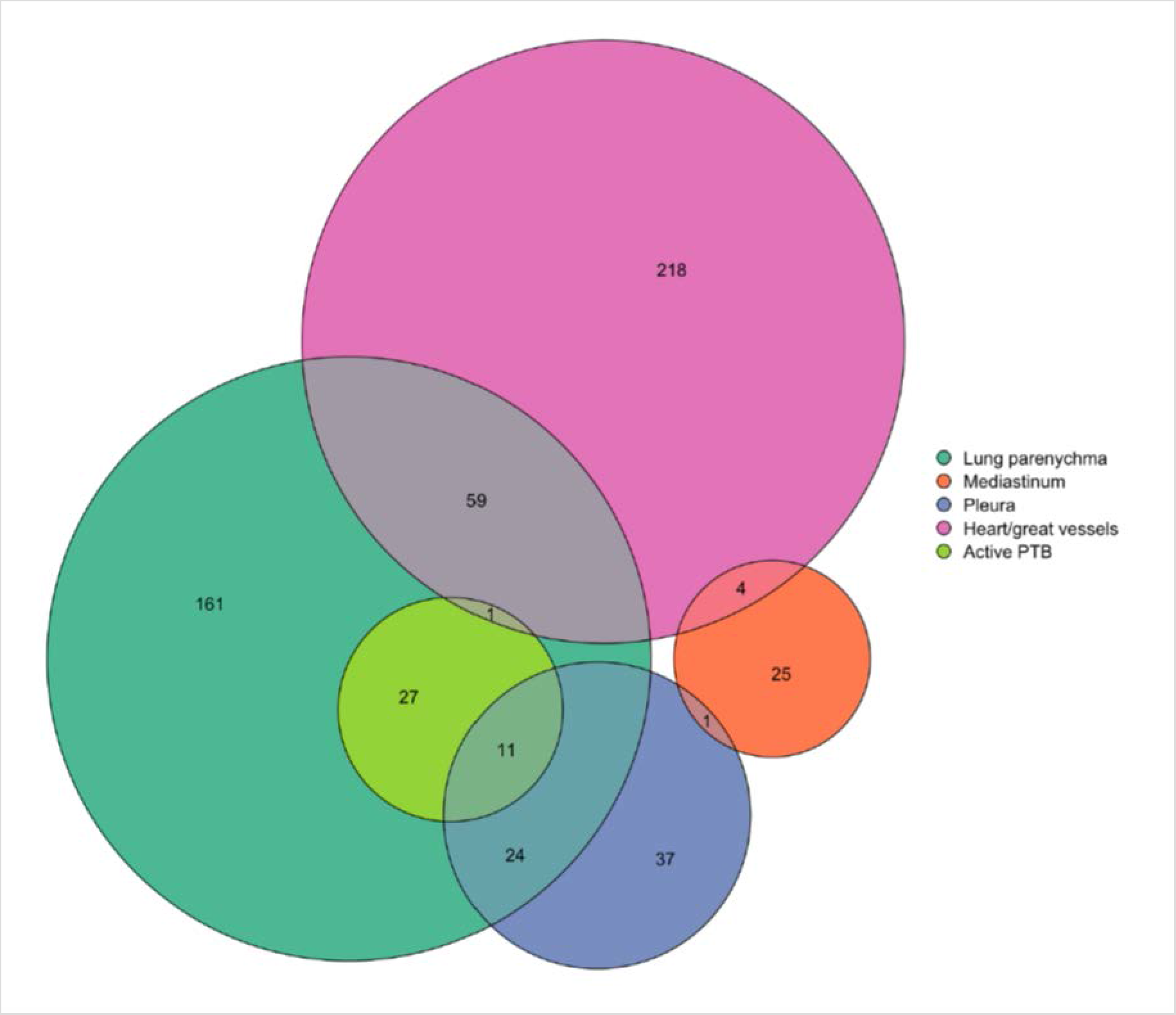
Euler diagram of abnormalities identified during the CXR study

Among the clinically relevant non-TB abnormalities, cardiomegaly was the most prevalent at 23.1% (95% CI

20.6%-25.6%), while cardiomegaly combined with features of cardiac failure occurred in 1.5% (95% CI 0.9%-2.4%). Non-specific patterns were noted in 4.6% (95% CI 3.5%-6.0%), while suspected chronic obstructive pulmonary disease (COPD), including emphysema, was present in 3.2% (95% CI 2.3%-4.4%). Mediastinal masses, excluding goitres, occurred in 0.8% (95% CI 0.4%-1.5%).

For TB related abnormalities, prevalence of minor post-TB lung changes, such as old/latent TB involving fewer than two lobes of damage/scarring was 7.6% (95% CI 6.1%-9.3%), active-TB was 3.6% (95% CI 2.6%-4.8%) and severe post-TB lung changes, i.e. bronchiectasis and/or destroyed lung, 1.4% (95% CI 0.8%-2.3%).

Between a quarter and a third of females had cardiomegaly (29%). For males 11.7% had cardiomegaly and 11.2% mild/moderate post TB lung changes. History of cough was a common feature across all diagnosis types with 69/250 (27.6%) of coughers having cardiomegaly. Out of the 40 participants with chest pain, 22.5 % (10.8-38.5%) had cardiomegaly and 20% (9.1-35.6%) had minor post-TB lung changes. Bacteriological confirmation of TB was found in all categories of reported TB-related lung abnormalities and 4/27, (14.8%, 4.2%-33.7%) of images reported as non-specific patterns (Table 2). Out of the 34 participants with a history of previous TB treatment, features consistent with minor post-TB lung changes were reported in 11(32.4%, 95% CI 17.4%-50.5%), active PTB in 6 (17.6%, 95% CI 6.8%-34.5%) and severe post-TB lung changes in 4 (11.8%, 95% CI 3.3%-27.5%).

**Table 2:** Prevalence and CAD4TB median scores of potentially clinically relevant radiological diagnoses with accompanying characteristics.

| RADIOLOGICAL DIAGNOSIS | Prevalence |  | CAD4TB scores | Sex |  | Age | History of cough | History of chest pain | Bacteriological TB confirmation (Xpert/Culture) |
| --- | --- | --- | --- | --- | --- | --- | --- | --- | --- |
|  | n=1123 | %, 95% CI |  | Female N=721, % (95%CI) | Male N=402, % (95%CI) |  |  |  |  |
| **Cardiomegaly** | 259 | 23.1% (20.6%-25.6%) | 50 (46-56) | 212/721, 29.4% (26.1%-32.9%) | 47/402, 11.7% (8.7%-15.2%) | 59(45-70) | 69/250, 27.6% (22.2%-33.6%) | 9/40, 22.5% (10.8%-38.5%) | 0/27, 0.0% (0.0%-12.8%) |
| **Cardiomegaly with heart failure** | 17 | 1.5% (0.9% - 2.4%) | 57(55-60) | 12/721, 1.7% (0.9% - 2.9%) | 5/402, 1.2% (0.4% - 2.9%) | 77(68-83) | 7/250, 2.8% (1.1% - 5.7%) | 0/40, 0.0% (0.0% - 8.8%) | 0/27, 0.0% (0.0%-12.8%) |
| **Suspected cardiac failure** | 21 | 1.9% (1.2% - 2.8%) | 57(53-60) | 16/721, 2.2% (1.3% - 3.6%) | 5/402, 1.2% (0.4% - 2.9%) | 75(67-83) | 8/250, 3.2% (1.4% - 6.2%) | 0/40, 0.0% (0.0% - 8.8%) | 0/27, 0.0% (0.0%-12.8%) |
| **Mild/moderate post-TB lung disease (*Old/latent TB with =< 2 lobe damage*)** | 85 | 7.6% (6.1% - 9.3%) | 67(58-79) | 40/721, 5.5% (4.0% - 7.5%) | 45/402, 11.2% (8.3%-14.7%) | 48(37-66) | 18/250, 7.2% (4.3%-11.1%) | 8/40, 20.0% (9.1%-35.6%) | 6/27, 22.2% (8.6%-42.3%) |
| **Non-specific opacification/interstitial pattern** | 52 | 4.6% (3.5% - 6.0%) | 57 (50-61) | 28/721, 3.9% (2.6% - 5.6%) | 24/402, 6.0% (3.9% - 8.8%) | 59(46-76) | 16/250, 6.4% (3.7%-10.2%) | 1/40, 2.5% (0.1%-13.2%) | 4/27, 14.8% (4.2%-33.7%) |
| **Active PTB** | 40 | 3.6% (2.6% - 4.8%) | 66 (55-72) | 15/721, 2.1% (1.2% - 3.4%) | 25/402, 6.2% (4.1% - 9.0%) | 36 (26-51) | 21/250, 8.4% (5.3%-12.6%) | 4/40, 10.0% (2.8%-23.7%) | 11/27, 40.7% (22.4%-61.2%) |
| **Suspected emphysema/COPD** | 36 | 3.2% (2.3% - 4.4%) | 57(52-61) | 16/721, 2.2% (1.3% - 3.6%) | 20/402, 5.0% (3.1% - 7.6%) | 68(50-73) | 11/250, 4.4% (2.2% - 7.7%) | 0/40, 0.0% (0.0% - 8.8%) | 0/27, 0.0% (0.0%-12.8%) |
| **Severe post-TB lung disease (*Bronchiectasis and or destroyed lung*)** | 16 | 1.4% (0.8% - 2.3%) | 76 (71-81) | 10/721, 1.4% (0.7% - 2.5%) | 6/402, 1.5% (0.5% - 3.2%) | 60(34-72) | 7/250, 2.8% (1.1% - 5.7%) | 0/40, 0.0% (0.0% - 8.8%) | 2/27, 7.4% (0.9%-24.3%) |
| **Mediastinal mass (excluding goitre /TB)** | 9 | 0.8% (0.4% - 1.5%) | 52 (47-59) | 7/721, 1.0% (0.4% - 2.0%) | 0/402, 0.5% (0.1% - 1.8%) | 61(56-71) | 2/250, 0.8% (0.1% - 2.9%) | 0/40, 0.0% (0.0% - 8.8%) | 0/27, 0.0% (0.0%-12.8%) |

#### CAD4TB analysis

The median score for all study CXRs was 50 (IQR 45–56.75) while for images classified as normal and abnormal by expert radiologists scores were 47 (IQR44-51) and 54 (IQR48-62) respectively. There was no difference between the sexes. The score for severe post-TB lung changes was highest at 76 (IQR 71-81). Active-TB and minor post-TB change scores were similar at 66 (IQR 55-73) and 67 (IQR58-79) respectively. The Xpert positive participants’ images had scores of 72 (IQR63-80). Abnormalities with lower scores included suspected COPD at 57 (IQR 52-61), non-specific patterns at 56 (IQR 50-61), mediastinal mass, excluding goitres, at 52 (IQR 47-59), cardiomegaly alone at 50 (IQR 46-56), and cardiomegaly with features of cardiac failure at 57 (IQR55-60).

### Discussion

The main finding from this analysis of X-ray images from the 2016 Kenya TB prevalence survey was that the use of CXR for TB population-based studies identified a large number of patients with abnormalities, including non-communicable diseases (NCDs) such as cardiovascular abnormalities and chronic respiratory diseases that require clinical attention. Clinically relevant cardiac and chronic pulmonary diseases accounted for 66% of the non-TB abnormalities in our setting.To our knowledge, this is the first study in sub-Saharan Africa to characterise and quantify non-TB CXR findings among participants who underwent mass screening as part of a population-based TB prevalence survey. TB prevalence surveys and ACF activities using mass CXR therefore offer opportunities to integrate disease screening efforts. The findings of our study are also timely in the wake of disruption in the health system caused by coronavirus disease (COVID-19).^22^ As countries accelerate ACF activities using CXR screening to make up for reductions in TB case-notification rates due to COVID-19, they could plan to integrate screening for other diseases.

At the outset of the study, we expected abnormalities to be primarily related to non-TB pulmonary disease. However, the most prevalent finding was cardiomegaly at 23.1% (95% CI 20.6%-25.6%). This is higher than in a smaller South African (n = 450) study, which reported cardiomegaly in 12.7% (95% C.I. 9.6%-15.8%) of CXRs taken during a vaccine study, among HIV-positive participants. Their participants had a much lower median age (34 vs 51 years) than in our study.^17^ Further, our study analysed images which had all originally been classified as abnormal. This, together with higher median age could explain our higher observed prevalence.

Calculation of the cardio-thoracic ratio (CTR) on CXR read by humans is a well-described affordable and reproducible screening method for cardiomegaly.^17, 23^ However, current studies, including our own, use CTR cut off values developed in Caucasian populations and there will be a need for robust validation of baseline CTR values for healthy populations in sub-Saharan Africa.^24^ CAD for detection of CTR is under development.^25^

In sub-Saharan Africa, the commonest causes of cardiomegaly are conditions of significant public health importance associated with premature mortality, including: hypertensive heart disease; cardiomyopathies; cor pulmonale; chronic rheumatic heart diseases; and ischaemic heart diseases.^23, 26^ Cardiomegaly has been associated with both higher body mass index (BMI) and higher median systolic blood pressure (BP).^17^ The high prevalence of cardiomegaly in our study supports exploration of the benefits of CVD screening during TB CXR screening as a potentially affordable public health intervention.^5, 23^ This could include adding relevant questions about previous hypertension diagnosis and treatment, measurement of BP and calculation of BMI. We recognize that a single BP reading is not diagnostic, but it could serve as an indicator for further follow-up.^27^ Health messaging on prevention of NCDs through recommendations on diet, such as reduction of commercial sugar and high salt diet, could be considered for integration in such programmes.^23^

Non-TB related respiratory pathology, including chronic respiratory diseases (CRD) were another significant finding in our cohort. In a Vancouver study in 1960s, three cases of significant previously unknown non-TB lung disease were identified for every new TB case; in our study, this figure was approximately 2:1.^6^ It should be noted that CXR alone has limited specificity for many of these conditions, especially in this cohort where very limited clinical information was available. The diagnoses of “non-specific airspace opacification” and “interstitial pattern” cover a range of possible pathologies, varying from incidental acute or chronic infective changes, not typical of TB, to non-infective pathology. COPD and emphysema cannot be diagnosed reliably on CXR alone, requiring spirometry and referral for further confirmation. However, CRD morbidity and mortality is on the rise, with the prevalence of COPD shown to range between 4%-25% in one systematic review in sub-Saharan Africa comparable to 3.6% (2.3%-4.4%) in our study.^28^ As expected, our study confirmed that screening for TB will detect alternative lung abnormalities in a significant number of non-TB cases and spirometry will be required for a subset of these patients.

Forty-four percent of the participants in our study with reported post-TB lung changes had a history of TB treatment. Among those, CXR revealed bronchiectasis and/or destroyed lung in 11% (95% CI 3.3%-27.5%), which is lower than reported in a prospective study in Malawi, that used computed tomography and reported > 40% bronchiectasis and 10% lobar destruction post-TB treatment.^21^ Bronchiectasis has a lower detection rate on CXR than CT, and will likely have been underestimated in our study. PTB is a risk factor for CRD and in the Malawi study ongoing clinical symptoms were associated with damage of three or more lobes,^21, 29^ which is comparable to the “destroyed lung” category in our CXR study. Unfortunately, patients with PTLD and chronic symptoms are likely to be treated empirically for recurrent TB.^30^ We therefore recommend that TB screening programs include protocols for PTLD management at primary care level to ensure patients are not unnecessarily retreated for TB. This also underscores the importance of bacteriological confirmation in patients with high CAD4TB scores. For those eligible, latent TB infection treatment should be offered.^31^ Our study also identified other less common findings for which interventions may be costly. For example, mediastinal masses that may represent lymphoma/ malignancy and would need referral for definitive diagnosis and management.

CAD4TB has been developed to rapidly identify people with CXR abnormalities indicative of TB.^18^ Our study had high median CAD4TB scores for all active PTB images at 66 (IQR 55-73) as defined by radiologist interpretation, as well as by bacteriological confirmation. Images with lower scores, including those with cardiomegaly (50, IQR 46-56) require review to ensure important non-TB pathology is detected. Analysis and modelling of the non-TB abnormalities CAD4TB scores is required. This will enable quantification of patients with non-TB pathology who would be flagged depending on various threshold cut-offs. This will translate to the numbers of patients with controllable NCDs missed per 100 000 population hence justifying further refinement of CAD algorithms to include non-TB diagnoses.

Our study used population-based national prevalence data and an explicit sampling approach to select images for review. Each image was read by two expert radiologists. Moderate inter-reader variability was mitigated by applying a third reader to resolve discrepancies. However, low specificity is an acknowledged issue with radiological classification. This was a retrospective study and we had limited clinical information available. HIV results were self-reported. Important information such as smoking history and pre-existing medical conditions were not collected during the survey. We were therefore not able to adequately correlate clinical symptoms or HIV serostatus with our findings. Though the prevalence survey protocol required those with other CXR abnormalities to be linked to a health facility within the cluster, we had no way of ascertaining if this was done, or obtaining data on final diagnosis and clinical outcome.

### Conclusion and recommendations

Our findings are strikingly similar to those of the 1940s study in Europe; that mass radiography can be used to tackle “fundamental problems of disease in the chest, both of the respiratory system and also of the heart” and “aid in detection of early and treatable non-TB disease”.^5^ Currently, TB screening activities using CXR and CAD software are focused on finding abnormalities consistent with TB.^16^ As countries embark on TB ACF activities, they need to be aware that other respiratory and non-respiratory pathologies are likely to be as, or more prevalent, than active TB. Mass screening with CXR therefore offers opportunity to screen for and address multiple important diseases.^5, 6^ Even though the algorithms or protocols for example in TB prevalence surveys do recommend that any other abnormalities should be referred as appropriate, there is no structured system for the detection and referral of such patients.^9, 15^ We recommend a patient-centered approach incorporating NCDs screening and health promotion during TB ACF activities. Clear referral pathways and follow-up plans for non-TB pathology could be incorporated during the planning of TB prevalence surveys and ACF activities.^6^ Prospective data collection about non-TB conditions identified during screening and economic impact could assist with health system planning. Our findings indicate a high prevalence of NCDs in the population. At primary care health facilities, prevention efforts for NCDs could be strengthened including health messaging, BP and BMI monitoring.

### Contributors

BNM and SBS were responsible for the study conceptualization. BNM, SBS, AO, EJ and PM designed the study. EM, JO and JS as key investigators of the Kenya prevalence survey, contributed in the study protocol development and approval for use of the data for this study. BNM, EJ, SBS, VM, BM, JO, EM and AO developed the study protocol. EJ, BNM, VM and BM developed study methodology, data collection tools and conducted the study. DK and RK developed the online reporting tool, conducted the sampling and managed the study data. PM conducted the data analysis and development of the figures. BNM, EJ, B Morton, SBS and PM played a major role in data interpretation and the writing of the manuscript. AO, B Morton, RO critically reviewed the manuscript. SBS was Director of the IMPALA Global Health Research Unit at the time of this work and played a major role in securing the funding for the work. As joint senior authors, SBS and PM provided final approval of the version to be published.

## Data Availability

The Kenya Division of National Tuberculosis, Leprosy and Lung Disease program is the custodian of the 2016 Kenya Tuberculosis Prevalence Survey data.

## Declaration of interest

We declare no competing interests.

## Data sharing

The Kenya National Tuberculosis, Leprosy and Lung Disease program is the custodian of the 2016 Kenya Tuberculosis Prevalence Survey data.

## Acknowledgements

We are grateful to the 2016 Kenya Tuberculosis Prevalence Survey team and the Division of National Tuberculosis, Leprosy and Lung Disease Program whose data we used for this secondary study. We would like to thank Jamilah Meghji and Jeremiah Chakaya who gave input during the conceptualisation of the study. We also acknowledge the team of expert radiologists engaged in the interpretation of the study X-ray images: Kenya team-Beatrice Mulama, Eva Maxine Angoro, Caroline Kebuka; UK team through Worldwide Radiology, UK: John Curtis, Alberto Alonso, Nigel Marchbank, Charlie Sayer, Laura Cormack and Ting Ting Zhang.

## Funding

This research was funded by the National Institute for Health Research (NIHR) (IMPALA, grant reference 16/136/35) using UK aid from the UK Government to support global health research. The views expressed in this publication are those of the author(s) and not necessarily those of the NIHR or the UK Department of Health and Social Care.

## Supplementary material

**Appendix 1:** Study radiologists details.

| Radiologist | Qualifications | Experience in TB imaging | Country | Reported |
| --- | --- | --- | --- | --- |
| 1 | Consultant radiologist) | 16 years | Kenya | Pilot study, Main study |
| 2 | Consultant radiologist | 19 years | Kenya | Pilot study, Main study |
| 3 | Consultant radiologist Fellowship in radio-oncology | 15 years | Kenya | Pilot study, Main study |
| 4 | Consultant radiologist | 10 years | Kenya | Main study |
| 5 | Consultant radiologist | 10 years | Kenya | Main study |
| 6 | Chest, Head and Neck radiologist | 3 years | United Kingdom | Main study |
| 7 | General Body Imaging Radiologist | 1 year | United Kingdom | Main study |
| 8 | Consultant Radiologist: Chest and Oncological Imaging | 30 years | United Kingdom | Main study |
| 9 | Specialist chest radiologist | 20 years | United Kingdom | Pilot study, Main study |
| 10 | Thoracic radiologist | 10 years | United Kingdom | Main study |
| 11 | Consultant radiologist | 15 years | United Kingdom | Pilot study, Main study-Third reader |
| 12 | Thoracic radiologist | 12 years | United Kingdom | Main study-Third reader |

**Appendix 2:**
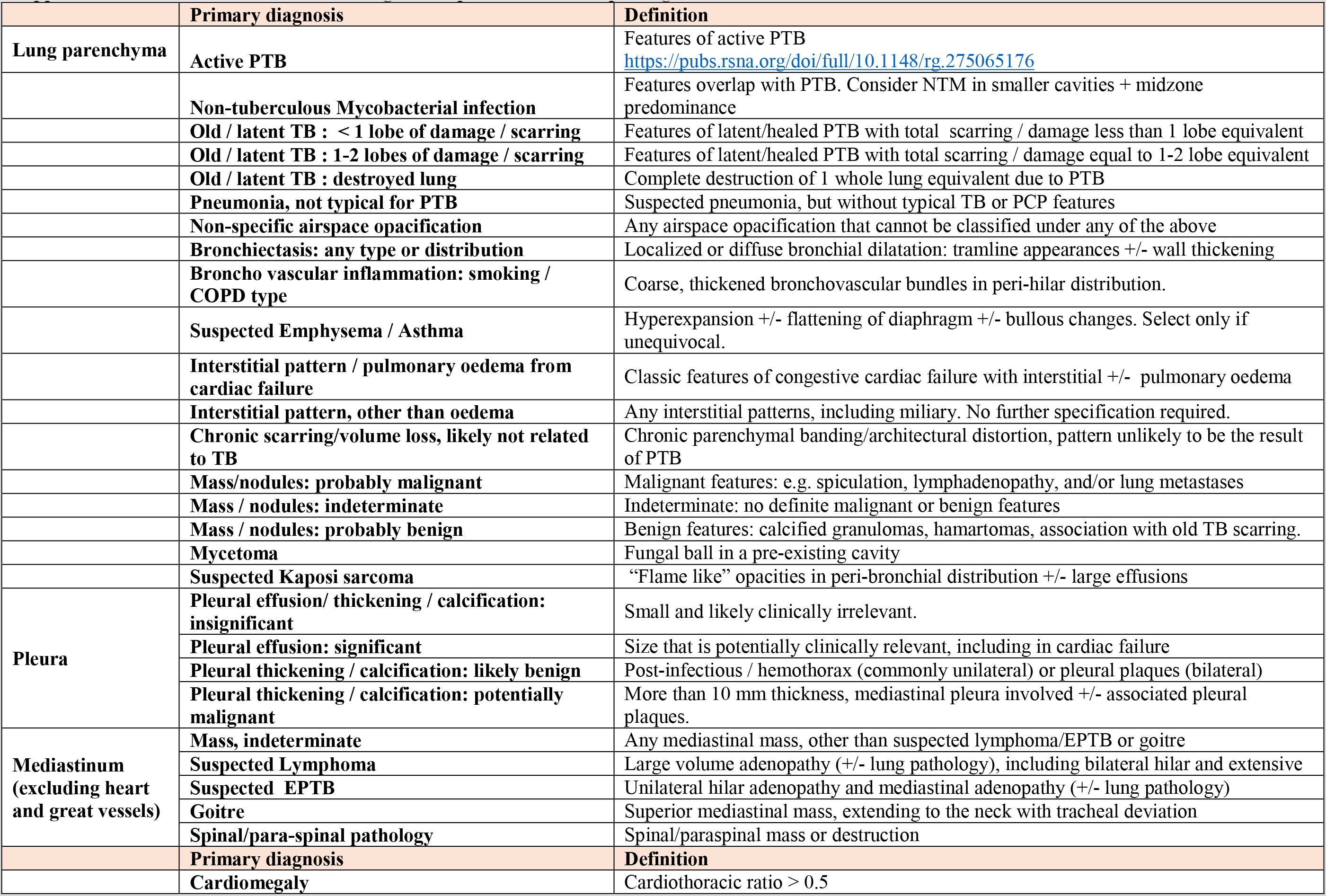
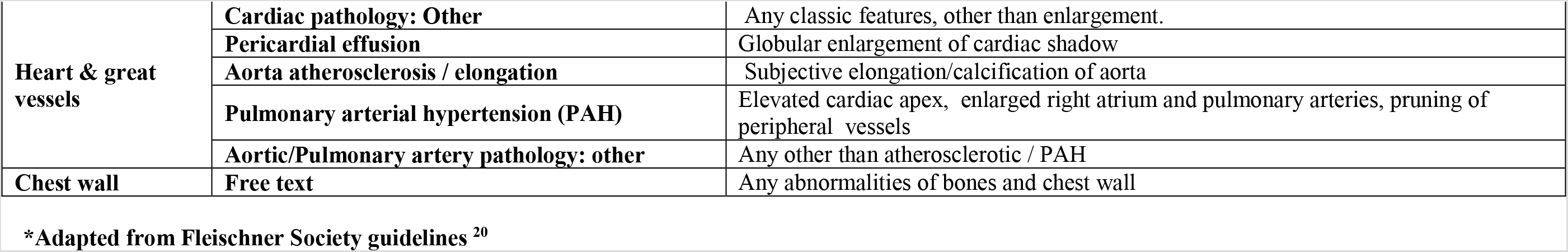
List of selected differential diagnostic options for CXR reporting and definitions*.

## Appendix 3

### PILOT STUDY

We conducted a pilot study from December 2018 to January 2019, to trial and refine a list of selected Chest X-ray diagnoses, to refine standard operating procedures for reporting and to estimate a sample size for the main study.

#### Pilot study methods

We developed an on-line, study specific CXR reporting tool which comprised four major diagnostic categories: lung parenchyma, heart and great vessels, and the pleura and mediastinum. Under each heading, a pick list of most common expected diagnoses was given, taking into consideration Kenyan disease epidemiology and prevalence survey cohort. During reporting of the CXRs, readers were required to select one or more primary diagnoses as participants could have more than one diagnosis., followed by the option of selecting up to two differential diagnoses. Due to limited specificity of CXR in many disease presentations, allowing for alternative diagnostic options was designed to capture those cases where a single, confident primary diagnosis could be not made. This study was aimed at deriving the prevalence of a final diagnosis rather than exploring image characteristics for each disease, the proforma did not capture detailed descriptions of each film.

Five radiologists were selected for the pilot: three consultant radiologists (MV, BM, BMM) from Kenya who had been part of the National TB Prevalence Survey team and two (JC, EJ) from the United Kingdom. Prior to the pilot readings, a test set of 20 X-rays was read independently by all radiologists, followed by discussion in a consensus meeting to ensure uniformity in reporting and application of the tool. Subsequently, each pilot image was read by a single radiologist only.

#### Pilot study sampling

We were unable to identify recent and relevant estimates from the literature to inform sample size estimates for prevalence of pulmonary abnormalities from similar settings in the pilot study. We therefore pragmatically set out to read 500 images (150 abnormal suggestive of TB; and 350 abnormal other) during the pilot, after which the detected prevalence of a set of predetermined pathologies was used to calculate a representative sample size for the main study. Once sampled, the labels of “Abnormal, suggestive of TB” and “Abnormal other” were removed to reduce the risk of bias.

#### Pilot study results

A total of 484 (97%) images were reported, 16 images were not read within the set time frame. The reporting was as follows: 288 (60%) as abnormal, 178 (37%) as normal, 8 (2.3%) were not interpretable. The abnormalities included: Heart and/or great vessel abnormalities 174 (37%), lung parenchyma abnormalities116 (24%), pleural abnormalities 39 (8.2%) and mediastinal abnormalities 8 (1.7%). In the images in the “abnormal other” category n = 344, cardiomegaly was most prevalent at 122 (36%) and great vessel abnormalities at 79 (23%). In the “abnormal suggestive of TB” category n = 122, old or latent TB was the most prevalent diagnosis at 17(13.9%) and active PTB at 16(13.1) %. Cardiomegaly in this category was at 9(7%).

**Appendix 4:**
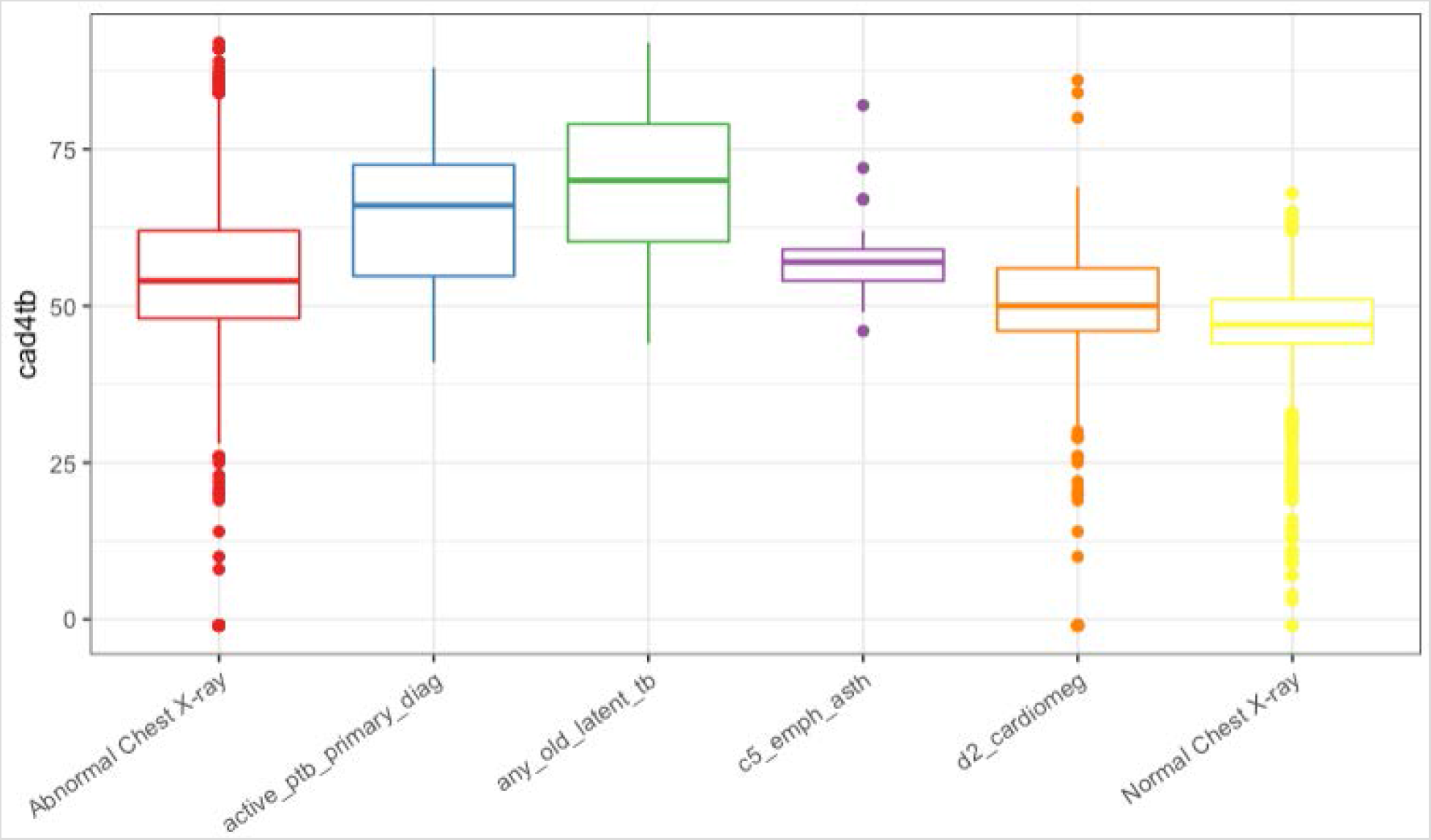
Box plot of CAD4TB scores for overall normal, abnormal and the clinically relevant diagnosis.

**Appendix 5:** Cohen’s Kappa scores for the expert radiologists inter-reader variability.

| Radiologists | A | B | C | D | E | F | G | H | I | J |
| --- | --- | --- | --- | --- | --- | --- | --- | --- | --- | --- |
| A | 1.000 | 0.372 | 0.178 | 0.000 | -0.421 | 0.623 | 0.438 | 0.292 | -0.429 | 0.016 |
| B | 0.372 | 1.000 | 0.659 | 0.322 | 0.459 | 0.700 | 0.475 | 0.114 | 0.503 | 0.214 |
| C | 0.178 | 0.659 | 1.000 | 0.046 | 0.409 | 0.814 | 0.215 | 0.204 | 0.298 | 0.224 |
| D | 0.000 | 0.322 | 0.046 | 1.000 | 0.765 | 0.500 | 0.488 | 0.678 | 0.232 | 0.500 |
| E | -0.421 | 0.459 | 0.409 | 0.765 | 1.000 | 0.698 | 0.576 | 0.487 | 0.422 | 0.688 |
| F | 0.623 | 0.700 | 0.814 | 0.500 | 0.698 | 1.000 | 0.036 | -0.227 | 0.529 | 0.800 |
| G | 0.438 | 0.475 | 0.215 | 0.488 | 0.576 | 0.036 | 1.000 | 0.175 | 0.455 | 0.500 |
| H | 0.292 | 0.114 | 0.204 | 0.678 | 0.487 | -0.227 | 0.175 | 1.000 | 0.427 | 0.157 |
| I | -0.429 | 0.503 | 0.298 | 0.232 | 0.422 | 0.529 | 0.455 | 0.427 | 1.000 | 0.620 |
| J | 0.016 | 0.214 | 0.224 | 0.500 | 0.688 | 0.800 | 0.500 | 0.157 | 0.620 | 1.000 |

## Notes

### Competing Interest Statement

The authors have declared no competing interest.

### Author Declarations

This study was conducted as part of the Kenya Prevalence survey ethics approval reference number SSC 2094 by Kenya Medical Research Institute.

